# Durability of SARS-CoV-2-specific IgG responses in saliva for up to 8 months after infection

**DOI:** 10.1101/2021.03.12.21252149

**Authors:** Pranay R. Randad, Nora Pisanic, Kate Kruczynski, Tyrone Howard, Magdielis Gregory Rivera, Kristoffer Spicer, Annukka A.R. Antar, Tristan Penson, David L. Thomas, Andrew Pekosz, Nelson Ndahiro, Lateef Aliyu, Michael J. Betenbaugh, Hannah Manley, Barbara Detrick, Morgan Katz, Sara Cosgrove, Clare Rock, Israel Zyskind, Jonathan I. Silverberg, Avi Z. Rosenberg, Priya Duggal, Yukari C. Manabe, Matthew H. Collins, Christopher D. Heaney

## Abstract

We evaluated the durability of IgG responses specific to SARS-CoV-2 nucleocapsid (N), receptor binding domain (RBD), and spike (S) antigens in saliva up to 8 months after RT-PCR-confirmed COVID-19 using a multiplex salivary assay. We estimated a half-life of 64 days (d) (95% CI: 49, 80 d) for N, 100 d for RBD (95% CI: 58, 141 d), and 148 d (95% CI: 62, 238 d) for S IgG responses in saliva, consistent with half-life estimates previously reported in blood. Saliva can serve as an alternative to blood to monitor humoral immune responses on a large scale following SARS-CoV-2 infection and vaccination for surveillance and assessment of population immunity.

## MAIN TEXT

There is a critical role for robust techniques to track humoral responses on a large scale following SARS-CoV-2 infection and vaccination for surveillance and assessment of population immunity. We previously developed and validated a multiplex bead-based immunoassay for detecting antibody responses to SARS-CoV-2 N, RBD, and S antigens in oral fluid (hereafter, saliva) .^1,2^ In this study, we apply this salivary assay to 531 saliva samples from 341 individuals up to 8 months after RT-PCR-confirmed SARS-CoV-2 infection to evaluate the durability of SARS-CoV-2 IgG responses. We hypothesized that the durability of SARS-CoV-2 N, RBD, and S IgG in saliva would exceed at least six months, comparable to those reported in blood. ^3–5^

SARS-CoV-2 N, RBD, and S IgG and the sum of their signal-to-cut off values (Σ[S/CO]) values peaked in saliva at ∼30 days post symptom onset (DPSO) (**Fig. 1a**). A linear fit was used to model the decay of SARS-CoV-2 N, RBD, and S IgG in saliva over time among saliva collected ≥30 DPSO (268 saliva samples from 237 people) (**Supplementary Table 1**).^4^ The estimated half-life was 64 days (d) (95% CI: 49, 80 d) for N Σ(S/CO), 100 d (CI: 58, 141 d) for RBD Σ(S/CO), and 148 d (95% CI: 59, 238 d) for S Σ(S/CO) (**Fig. 1a, Supplementary Table 2**). RBD and S Σ(S/CO) were more stable over time compared to N. These half-life estimates for salivary IgG are similar with those reported for circulating antibodies in plasma: 67 d (95% CI: 49, 105 d) for N IgG, 83 d (95% CI: 62, 127 d) for RBD IgG, and140 d (95% CI:89, 329 d) for S IgG.^4^

**Figure 1.**
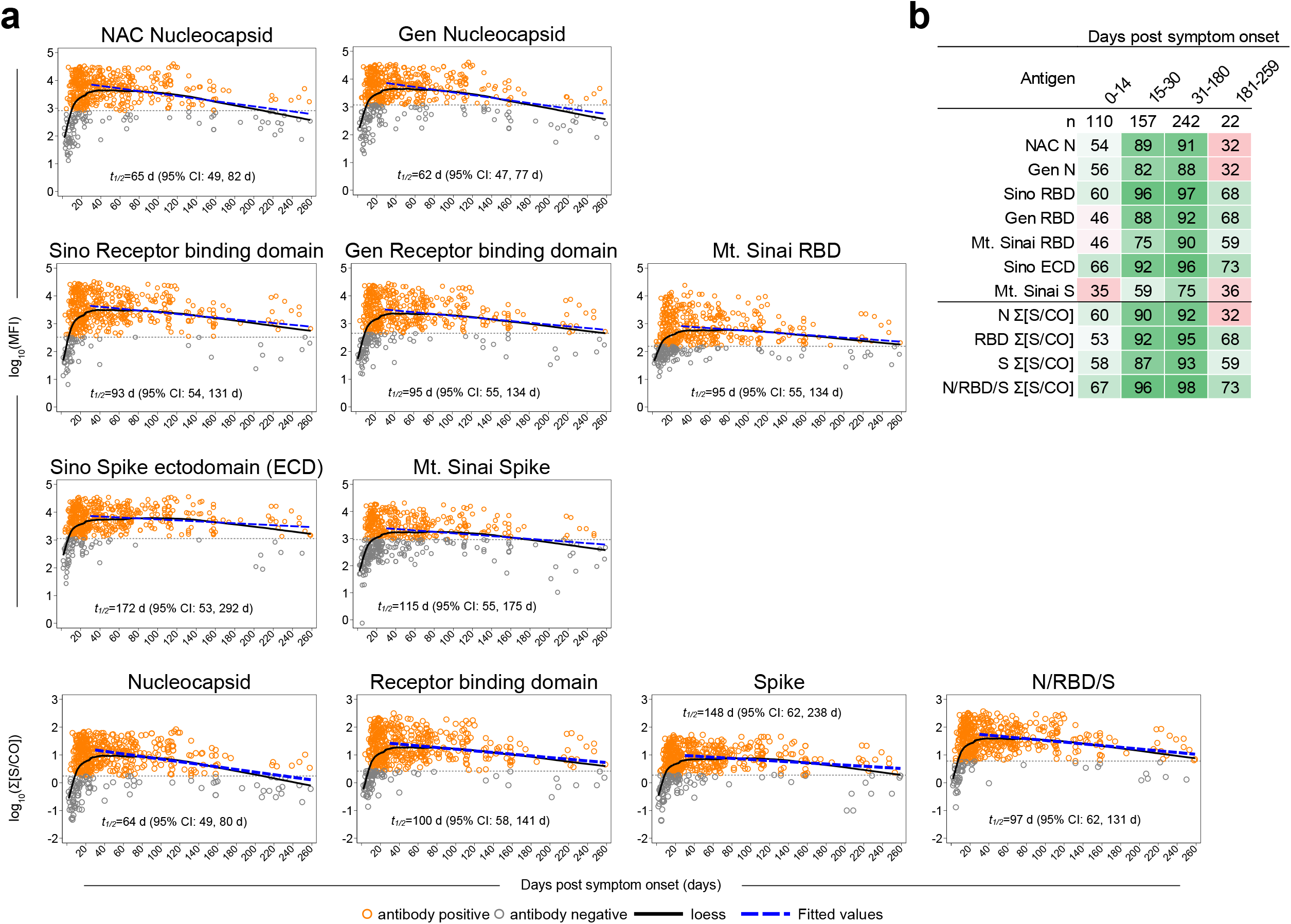
SARS-CoV-2-specific IgG responses in saliva over time. **(a)** Log_10_median fluorescence intensity (MFI) of SARS-CoV-2-specific IgG responses to nucleocapsid (N), receptor binding domain (RBD), spike (S), and Σ[S/CO] values among n=531 saliva samples (from n=341 COVID-19 cases) over time. The solid black line represents temporal kinetics (estimated by loess spline). Dashed blue line represents the estimated slope with half-life (*t*_*1/2*_) and 95% confidence interval using a linear fit model (n=268 saliva samples from 237 cases). Dashed gray lines indicate cut-off values. Hollow orange and gray circles represent saliva samples classified as positive or negative, respectively. **(b)** Heat map detailing the proportion of saliva samples classified as positive by days post symptom onset. *Note*. Log_10_ data is shown for all plots; Σ[S/CO], sum of signal to cut-off; NAC, Native Antigen Company; Gen, GenScript; Sino, SinoBiological.

Most saliva samples from COVID-19 subjects were positive for SARS-CoV-2 N (223/242; 92%), RBD (231/242; 95%), and S (224/242; 93%) IgG between 1-6 months post symptom onset (31-180 d) (**Fig. 1b**). The proportion of saliva samples positive for N, RBD, and S IgG from 6-8 months post symptom onset (181-259 d) was 32% (7/22), 68% (15/22) and 59% (13/22), respectively. Using the combined N/RBD/S IgG Σ[S/CO], positivity for SARS-CoV-2 IgG was 98% (236/242; 117 from females and 122 from males) between 1-6 months, and 73% (16/22; 10 from females and 12 from males) between 6-8 months (**Fig. 1b**), indicating that most COVID-19 subjects are positive for SARS-CoV-2 IgG in saliva up to 8 months post symptom onset and that incorporating the IgG response to multiple SARS-CoV-2 antigens can improve the sensitivity of saliva-based SARS-CoV-2 antibody testing.

There was evidence of a sex-specific interaction for N IgG and Σ[S/CO], with males having more durable N IgG levels in saliva over time compared to females (n=265 saliva samples, 128 from 112 females and 137 from 122 males) (**Fig. 2a, Supplementary Table 3 and 4**). In general, half –life estimates for N, RBD, and S IgG in saliva were longer in males than females (**Supplementary Table 3**). We detected a trend of elevated RBD IgG and Σ[S/CO] in saliva of males compared to females (**Supplementary Table 5**), consistent with previous reports in blood.^4,6,7^

**Figure 2.**
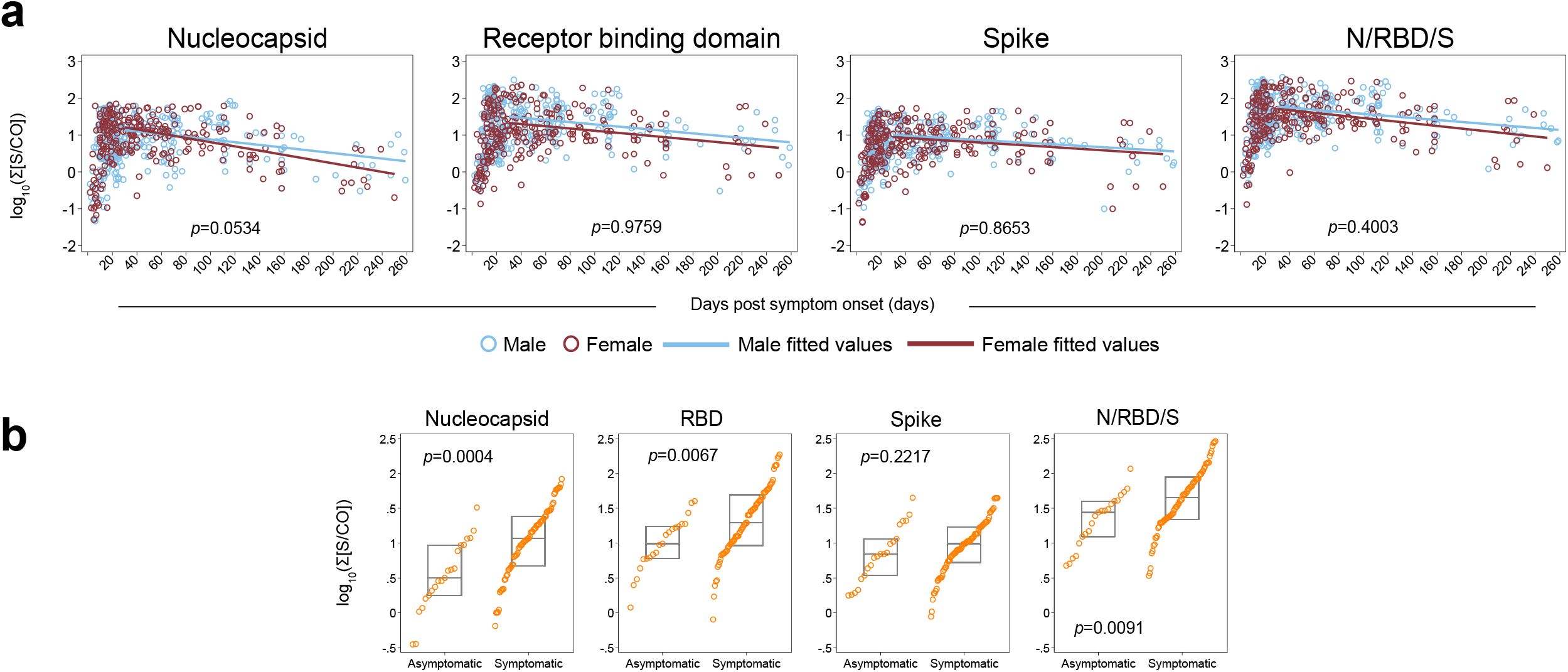
SARS-CoV-2-specific IgG responses in saliva over time, by sex and COVID-19 symptoms status. (**a**) Nucleocapsid (N) IgG Σ[S/CO], receptor binding domain (RBD) IgG Σ[S/CO], spike (S) Σ[S/CO], and N/RBD/S Σ[S/CO] in saliva over time, by sex (n=528 saliva samples, 234 from 127 females and 294 from 211 males). Hollow light-blue and maroon circles represent males and females, respectively. Light-blue and maroon lines represent the linear fit for males and females, respectively (n=265 saliva samples, 128 from 112 females and 137 from 122 males). *p* values were estimated via likelihood-ratio test. (**b**) N IgG Σ[S/CO], RBD IgG Σ[S/CO], S Σ[S/CO], and N/RBD/S Σ[S/CO] in saliva by symptoms (93 saliva samples from 93 COVID-19 cases, 21 asymptomatic and 72 symptomatic, ranging from 72-174 DPSO). *p* values represent comparison of medians between groups via Wilcoxon-Mann-Whitney test. Box gray lines represent 25^th^, 50^th^, and 75^th^ percentile. *Note*. Log_10_ data is shown for all plots; Σ[S/CO], sum of signal to cut-off; NAC, Native Antigen Company; Gen, GenScript; Sino, Sino Biological.

We observed elevated median N and RBD IgG and Σ[S/CO] values among symptomatic compared to asymptomatic COVID-19 subjects (**Fig. 2b, Supplementary Table 6**), suggesting that asymptomatic subjects may be less likely to have durable IgG responses in saliva. In this analysis, we restricted the number of samples used from symptomatic subjects to better match the more limited set available from asymptomatic subjects (93 total saliva samples analyzed from 93 subjects, 21 asymptomatic and 72 symptomatic, from 72-174 DPSO). These results are consistent with reports of elevated SARS-CoV-2 antibodies in blood among individuals with moderate/severe compared to asymptomatic/mild COVID-19.^4,5^ A larger set of saliva samples from asymptomatic COVID-19 subjects should be tested in future studies.

We chose to conduct a cross-sectional analysis to achieve a large sample size and include time points 6-8 months post symptom onset. Ideally, this analysis would be repeated in a large, longitudinal sample set should it come available. Strengths of this analysis include saliva samples ranging from 2-259 DPSO, allowing for analysis of durability for up to 8 months, and the use of optimized algorithms for analyzing SARS-CoV-2 IgG multiplex data in saliva.

These results demonstrate that SARS-COV-2 RBD and S IgG are durable in saliva for up to 8 months post symptom onset. Durability appears to be increased among males versus females and symptomatic vs. asymptomatic SARS-CoV-2 infections. These results are consistent with what has been previously reported in blood ^3,4^. Overall, these results support the utility of saliva as an accurate and non-invasive alternative to blood for longitudinal SARS-CoV-2 antibody testing and large-scale surveillance.

## METHODS

Saliva samples were collected from participants enrolled into five groups of cohorts, which are described in the **Supplementary Text**, along with methods for saliva sample collection, processing, testing, and analysis for SARS-CoV-2 IgG in saliva using a 23-plex bead-based multiplex assay.

We calculated the median fluorescence intensity (MFI) signal-to-cutoff (S/CO) for seven individual SARS-CoV-2 antigens, using MFI cut-off values derived using a set of pre-pandemic saliva samples collected before January 2019 (n=265 saliva samples) (**Supplementary Table 7**). The Σ[S/CO] for two N antigens (NAC N and Gen N), three RBD antigens (Sino RBD, Gen RBD and Mt. Sinai RBD), two S antigens (Sino ECD and Mt. Sinai S), and all seven antigens (N/RBD/S) were used to estimate SARS-CoV-2 IgG durability in saliva over time. Antibody durability was also analyzed with each antigen separately using blank subtracted MFI values. Saliva samples with <15ug/ml of total IgG were excluded from analysis. Loess splines were used to visualize the kinetics of IgG in saliva over DPSO. The RT-PCR test date was used if symptom onset data was not available. To allow for comparability with previous studies in blood,^4^ a generalized linear model, clustered on individual to account for multiple samples, was used to model the durability of SARS-CoV-2 IgG in saliva. Log_2_ transformed data was used to estimate IgG half-life (−1/β coefficient).^4^ For sex analysis, interaction was tested using a likelihood-ratio test. A generalized linear model adjusted for DPSO and clustered on individual was used to compare SARS-CoV-2 IgG levels by sex. A Wilcoxon-Mann-Whitney test was used to compare SARS-CoV-2 IgG levels by symptoms.

## Data Availability

The datasets generated during and/or analyzed during the current study are available from the corresponding author on reasonable request.

## ACKNOWLEDGEMENTS

Funding was provided by the Johns Hopkins COVID-19 Research and Response Program, the FIA Foundation, a gift from the GRACE Communications Foundation (**C.D.H**., **P.R.R**., **N.P**., **K.K.)**, NIAID grants R21AI139784 (**C.D.H**. and **N.P**), National Institute of Environmental Health Sciences (NIEHS) grant R01ES026973 (**C.D.H**., **N.P**., **K.K**.), NIAID grant R01AI130066 and NIH grant U24OD023382 (**C.D.H**), CDC epicenter grant (**C.R**., **M.K**., **S.C**.), NIAID grant 3R01AI148049 (**P.D**., **D.L.T**.), the Johns Hopkins University School of Medicine COVID-19 Research Fund, the Sherrilyn and Ken Fisher Center for Environmental Infectious Diseases Discovery Program, and NIH grants U54EB007958-12, U5411090366, U54HL143541-02S2, and UM1AI068613 (**Y.C.M**.), National Institute for Innovation in Manufacturing Biopharmaceuticals (NIIMBL) and U.S. U.S. Department of commerce, NIST award 70NANB20H037 (**M.B**., **N.N**., **L.A**.), and NIH/NIAID Center of Excellence in Influenza Research and Surveillance contract HHS N2772201400007C (**A.P**.). The funders had no role in study design, data analysis, decision to publish, or preparation of the manuscript.

## AUTHOR CONTRIBUTIONS

All authors reviewed and edited all sections of the article. P.R.R. conducted all data analysis and developed the manuscript. N.P. generated data, optimized data analysis algorithms and curated the data set. K.K. handled laboratory logistics and generated data. A.Z.R., I.Z., J.I.S., M.K., P.D., Y.C.M., and M.H.C. led and coordinated specimen collection efforts. M.H.C. and C.D.H. developed the project concept.

