## Supplementary Material for "Durability of SARS-CoV-2-specific IgG responses in saliva for up to 8 months after infection"

### Supplementary Text

Saliva samples were collected from participants enrolled into five groups of cohorts: **1**) collaborators from Emory University provided 29 saliva samples from 29 patients enrolled in one of two studies[1,2]: (i) PCR-confirmed COVID-19 cases were recruited for sample donation during acute illness (all hospitalized) and/or during convalescence (some subjects were previously hospitalized, others were not). Convalescent time points were not synchronized but most were completed in the first 1-2 months post symptom onset (Emory IRB#00022371) (ii) patients with symptoms consistent with COVID-19 presenting to an ambulatory testing center were recruited to donated convalescent specimens (typically 21-56 days post symptom onset). Results of clinical PCR testing were obtained for all participants, with ~20% of this group being diagnosed with COVID-19 (Emory IRB#00110683); **2**) collaborators from Johns Hopkins provided 292 saliva samples from 102 individuals presenting at one of the Johns Hopkins Health System COVID-19 testing sites and testing positive for SARS-CoV-2 by RT-PCR;[3] **3**) collaborators from Johns Hopkins provided 27 saliva samples collected from 27 residents and staff with documented history of nasopharyngeal swabs testing positive by RT-PCR from three skilled nursing facilities in Maryland with prior COVID-19 outbreaks. Resident saliva samples were collected by trained research staff on-site at the facility and staff saliva samples were self-collected. Symptoms were obtained from the nursing home staff and residents and documented by the Johns Hopkins Testing Team at the time of PCR testing[4]; **4**) Saliva samples (n=157) from 99 PCR-positive adults adults and 58 minors (unpublished) who were recruited by local not-for-profit and social service organizations within orthodox Jewish communities after recovering from their acute illness were provided by the MITZVA cohort[5]; and **5**) collaborators from Johns Hopkins provided 23 saliva samples from 23 ambulatory patients that tested positive for SARS-CoV-2 infection by RT-PCR at Johns Hopkins in March of 2020 enrolled in a host genetics study of COVID-19 (unpublished).

Individuals were classified as COVID-19 cases (n=531 saliva samples from 341 individuals) if they were confirmed to be infected with SARS-COV-2 by RT-PCR assays on nasal and pharyngeal swab specimens. Saliva samples were either self-collected at home (n=236) or self-collected at a research site (n=295) (**Supplementary Table 1**). Saliva samples were collected by instructing participants to gently brush their gum line for 1-2 mins, or until saturated, with an Oracol S14 saliva collection device (Malvern Medical Developments, UK). This saliva collection method specifically harvests gingival crevicular fluid enriched with primarily IgG antibodies derived from passive leakage of serum into the oral compartment[6]. The saturated sponge was then inserted into the storage tube, capped, and stored at 4°C, whenever possible, until processing. Saliva was separated from the Oracol S14 sponge by centrifugation (10 min at 1,500 × *g*) and transferred into the attached 2-ml cryovial. Saliva samples were heat inactivated at 60ºC or 56ºC and stored at -80ºC until use.

Saliva was tested for IgG binding to SARS-CoV-2 and additional virus antigens using a 23-plex bead-based multiplex assay.[1] Saliva (10ul) was added to 40ul of assay buffer (PBST with 1% BSA) and 1,000 coupled beads/bead-set in each well. Assay buffer was used instead of saliva in one to two wells/plate for background fluorescence subtraction. SARS-CoV-2 IgG positive saliva with IgG signal was spiked into pre-pandemic negative saliva and used as a positive control. The same pre-pandemic saliva was used as negative control. Phycoerythrin-labeled anti-human IgG diluted 1:100 in assay buffer was used to detect IgG signal in the saliva. The plate was read on a Luminex MAGPIX instrument. The total IgG concentration in saliva was determined using Salimetrics Salivary Human Total IgG ELISA Kits according to the manufacturer’s instructions, with incubation times reduced to 1 hour (approved by manufacturer).

**Supplementary Table 1. Participant characteristics.**

| | COVID-19 cases<br>n=341 | Saliva samples<br>n=531 |
|---|---|---|
| | n (%) | n (%) |
| Age (mean [SD]; range) | 48 (18); 14-90 | |
| Sex | | |
| Female | 127 (37.2) | 234 (43.5) |
| Male | 211 (61.9) | 294 (55.4) |
| Missing | 3 (0.88) | 3 (0.56) |
| Symptoms | | |
| Asymptomatic | 23 (6.74) | 23 (4.33) |
| Symptomatic | 314 (92.1) | 504 (94.9) |
| Missing | 4 (1.17) | 4 (0.75) |
| Days post symptom onset (range) (n=531)* | 2-259 | |
| <30 days | 148 (43.4) | 263 (49.5) |
| ≥30 days | 237 (69.5) | 268 (50.5) |
| Collection location[†] | | |
| Clinic | 295 (86.5) | 295 (55.56) |
| Home | 96 (28.2) | 236 (44.44) |
| Saliva collection frequency | | |
| Single time point | 275 (80.6) | 275 (51.8) |
| Multiple time points | 66 (19.4) | 256 (48.2) |

*44 individuals contributed saliva samples collected at <30 days and ≥30 days; days post nucleic acid amplification test result was used when symptom onset data was not available

[†]50 individuals contributed saliva samples collected at the clinic and home

**Supplementary Table 2. SARS-CoV-2 IgG antibody half-life estimates in saliva.***

| Antigen | N | Model | β coefficient (95% CI)† | $p$ value† | $t_{1/2}$ (95% CI) ‡ | $p$ value‡ |
|---|---|---|---|---|---|---|
| NAC N | 268 | Linear | -0.004 (-0.006, -0.003) | 0.0001 | 65d (49, 82d) | 0.0001 |
| Gen N | 268 | Linear | -0.004 (-0.006, -0.004) | 0.0001 | 62d (47, 77d) | 0.0001 |
| Sino RBD | 268 | Linear | -0.003 ( -0.005, -0.002) | 0.0001 | 93d (54, 131d) | 0.0001 |
| Gen RBD | 268 | Linear | -0.003 ( -0.004, -0.002) | 0.0001 | 95d (55, 134d) | 0.0001 |
| Mt. Sinai RBD | 268 | Linear | -0.002 (-0.004, -0.001) | 0.0001 | 124d (62, 186d) | 0.0001 |
| Sino ECD | 268 | Linear | -0.002 (-0.003, -0.001) | 0.005 | 172d (53, 292d) | 0.005 |
| Mt. Sinai S | 268 | Linear | -0.003 (-0.004, -0.001) | 0.0001 | 115d (55, 175d) | 0.0001 |
| N Σ[S/CO] | 268 | Linear | -0.005 (-0.006, -0.004) | 0.0001 | 64d (49, 80d) | 0.0001 |
| RBD Σ[S/CO] | 268 | Linear | -0.003 (-0.004, -0.002) | 0.0001 | 100d (58, 141d) | 0.0001 |
| S Σ[S/CO] | 268 | Linear | -0.002 (-0.003, -0.001) | 0.001 | 148d (59, 238d) | 0.001 |
| N/RBD/S Σ[S/CO] | 268 | Linear | -0.003 (-0.004, -0.002) | 0.0001 | 97d (62, 131d) | 0.0001 |

**Note*: N, nucleocapsid; RBD, receptor binding domain; S, spike; ECD, Spike ectodomain; Σ[S/CO], sum of signal to cut-off ratio; Gen, GenScript; NAC, Native Antigen Company; Sino, Sino Biological; $t_{1/2}$ IgG antibody half-life in saliva.

†β coefficients and $p$ value estimated using a generalized linear model and $\log_{10}$ data, clustered on individual.

‡Antibody half-life and $p$ value estimated using a generalized linear model and log2 data, clustered on individual.

**Supplementary Table 3. SARS-CoV-2 antigen-specific IgG antibody half-life estimates in saliva, by sex.***

| Antigen | | N | Model | beta coefficient (95% CI) † | *p* value† | t1/2 (95% CI) ‡ | *p* value‡ |
|---|---|---|---|---|---|---|---|
| NAC N | | | | | | | |
| | Female | 128 | Linear | -0.006 (-0.007, -0.004) | 0.0001 | 54d (38, 69d) | 0.0001 |
| | Male | 137 | Linear | -0.004 (-0.005, -0.002) | 0.0001 | 82d (47, 118d) | 0.0001 |
| Gen N | | | | | | | |
| | Female | 128 | Linear | -0.006 (-0.008, -0.004) | 0.0001 | 50d (35, 64d) | 0.0001 |
| | Male | 137 | Linear | -0.004 (-0.005, -0.002) | 0.0001 | 81d (47, 115d) | 0.0001 |
| Sino RBD | | | | | | | |
| | Female | 128 | Linear | -0.003 (-0.006, -0.001) | 0.004 | 89d (29, 149d) | 0.004 |
| | Male | 137 | Linear | -0.003 (-0.005, -0.002) | 0.0001 | 93d (47, 139d) | 0.0001 |
| Gen RBD | | | | | | | |
| | Female | 128 | Linear | -0.003 (-0.005, -0.001) | 0.005 | 92d (30, 155d) | 0.004 |
| | Male | 137 | Linear | -0.003 (-0.005, -0.002) | 0.0001 | 94d (48, 140d) | 0.0001 |
| Mt. Sinai RBD | | | | | | | |
| | Female | 128 | Linear | -0.002 (-0.004, -0.001) | 0.014 | 120d (26, 215d) | 0.012 |
| | Male | 137 | Linear | -0.002 ( -0.004, -0.001) | 0.003 | 126d (43, 208d) | 0.003 |
| ECD | | | | | | | |
| | Female | 128 | Linear | -0.002 (-0.004, 0.0003) | 0.091 | 159d (-24, 342d) | 0.089 |
| | Male | 137 | Linear | -0.002 ( -0.003, -0.0002) | 0.018 | 184d (34, 335d) | 0.016 |
| Mt. Sinai S | | | | | | | |
| | Female | 128 | Linear | -0.003 (-0.005, -0.0003) | 0.028 | 108d (13, 203d) | 0.026 |
| | Male | 137 | Linear | -0.002 (-0.004, -0.001) | 0.001 | 123d (49, 197d) | 0.001 |
| N Σ[S/CO] | | | | | | | |
| | Female | 128 | Linear | -0.006 (-0.007, -0.004) | 0.0001 | 52d (37, 69d) | 0.0001 |
| | Male | 137 | Linear | -0.004 (-0.005, -0.002) | 0.0001 | 82d (48, 117d) | 0.0001 |
| RBD Σ[S/CO] | | | | | | | |
| | Female | 128 | Linear | -0.003 (-0.005, -0.001) | 0.005 | 98d (31, 164d) | 0.004 |
| | Male | 137 | Linear | -0.003 (-0.005, -0.002) | 0.0001 | 99d (49, 148d) | 0.0001 |
| S Σ[S/CO] | | | | | | | |
| | Female | 128 | Linear | -0.002 (-0.004, 0.0001) | 0.062 | 143d (-5, 291d) | 0.059 |
| | Male | 137 | Linear | -0.002 (-0.003, -0.0006) | 0.005 | 155d (46, 265d) | 0.005 |
| N/RBD/S Σ[S/CO] | | | | | | | |
| | Female | 128 | Linear | -0.004 (-0.006, -0.002) | 0.0001 | 84d (39, 130d) | 0.0001 |
| | Male | 137 | Linear | -0.003 (-0.004, -0.001) | 0.0001 | 110d (59, 161d) | 0.0001 |

**Note*: N, nucleocapsid; RBD, receptor binding domain; S, spike; ECD, Spike ectodomain; Σ[S/CO], sum of signal to cut-off ratio; Gen, GenScript; NAC, Native Antigen Company; Sino, Sino Biological; $t_{1/2}$ IgG antibody half-life in saliva.

†β coefficients and *p* value estimated using a generalized linear model and $log_{10}$ data, clustered on individual

‡Antibody half-life and *p* value estimated using a generalized linear model and log2 data, clustered on individual

**Supplementary Table 4. Tests of homogeneity of the association between days post-COVID-19 symptom onset and each SARS-CoV-specific IgG response, by sex.***

| Antigen | N | Chi-square value (df)† | *p* value |
|---|---|---|---|
| NAC N | 265 | 3.05 (1) | 0.0809 |
| Gen N | 265 | 4.55 (1) | 0.0329 |
| Sino RBD | 265 | 0.01 (1) | 0.9075 |
| Gen RBD | 265 | 0.00 (1) | 0.9621 |
| Mt. Sinai RBD | 265 | 0.01 (1) | 0.9292 |
| Sino ECD | 265 | 0.07 (1) | 0.7900 |
| Mt. Sinai S | 265 | 0.09 (1) | 0.7623 |
| N Σ[S/CO] | 265 | 3.73 (1) | 0.0534 |
| RBD Σ[S/CO] | 265 | 0.00 (1) | 0.9759 |
| S Σ[S/CO] | 265 | 0.03 (1) | 0.8653 |
| N/RBD/S Σ[S/CO] | 265 | 0.71 (1) | 0.4003 |

**Note*: N, nucleocapsid; RBD, receptor binding domain; S, spike; ECD, Spike ectodomain; Σ[S/CO], sum of signal to cut-off ratio; Gen, GenScript; NAC, Native Antigen Company; Sino, Sino Biological.

†Likelihood-ratio test; df, degrees of freedom.

**Supplementary Table 5. Association between sex and SARS-CoV-2 IgG antibody levels in saliva.***

| Antigen | N | β coefficient (95% CI)† | *p* value† |
|---|---|---|---|
| NAC N | 265 | 0.046 (-0.087, 0.179) | 0.4980 |
| Gen N | 265 | 0.036 (-0.096, 0.168) | 0.5900 |
| Sino RBD | 265 | 0.195 (0.066, 0.324) | 0.0030 |
| Gen RBD | 265 | 0.184 (0.051, 0.318) | 0.0070 |
| Mt. Sinai RBD | 265 | 0.099 (-0.038, 0.236) | 0.1570 |
| Sino ECD | 265 | 0.044 (-0.071, 0.158) | 0.4540 |
| Mt. Sinai S | 265 | 0.138 (0.007, 0.270) | 0.0400 |
| N Σ[S/CO] | 265 | 0.040 (-0.090, 0.171) | 0.5470 |
| RBD Σ[S/CO] | 265 | 0.171 (0.042, 0.299) | 0.0090 |
| S Σ[S/CO] | 265 | 0.066 (-0.049, 0.181) | 0.2630 |
| N/RBD/S Σ[S/CO] | 265 | 0.104 (-0.007, 0.214) | 0.0650 |

*Note*: N, nucleocapsid; RBD, receptor binding domain; S, spike; ECD, Spike ectodomain; Σ[S/CO], sum of signal to cut-off ratio; Gen, GenScript; NAC, Native Antigen Company; Sino, Sino Biological.

†β coefficients and *p* values were estimated using a generalized linear model with log10 SARS-CoV-2 antigen-specific IgG as the dependent variable and sex as the independent variable (females as referent category), adjusted for days post symptom onset and clustered on individual.

**Supplementary Table 6. SARS-CoV-2 antigen-specific IgG median fluorescence intensity (MFI) in saliva by COVID-19 symptom status.***

| | Asymptomatic | | | | | | Symptomatic | | | | | | |
|---|---|---|---|---|---|---|---|---|---|---|---|---|---|
| Antigen | n | Min | Max | median | mean | std. dev | n | Min | Max | median | mean | std. dev | *p* value† |
| NAC N | 21 | 168 | 14577 | 1747 | 2890 | 3288 | 72 | 284 | 39553 | 5774 | 9105 | 9270 | 0.0004 |
| Gen N | 21 | 164 | 16840 | 1416 | 3033 | 3901 | 72 | 311 | 40554 | 5636 | 8847 | 9130 | 0.0005 |
| Sino RBD | 21 | 165 | 6674 | 1796 | 2160 | 1732 | 72 | 63 | 26297 | 3361 | 6098 | 6384 | 0.0023 |
| Gen RBD | 21 | 138 | 4799 | 1190 | 1555 | 1214 | 72 | 64 | 21907 | 2393 | 4603 | 5238 | 0.0058 |
| Mt. Sinai RBD | 21 | 60 | 1826 | 332 | 544 | 487 | 72 | 54 | 9664 | 653 | 1341 | 1968 | 0.0525 |
| Sino ECD | 21 | 1598 | 35462 | 5560 | 8795 | 8766 | 72 | 646 | 34005 | 8424 | 10073 | 8011 | 0.2823 |
| Mt. Sinai S | 21 | 318 | 12138 | 1693 | 2397 | 2788 | 72 | 54 | 17350 | 2244 | 3396 | 3639 | 0.1544 |
| N Σ[S/CO] | 21 | 0.4 | 32.3 | 3.2 | 6.2 | 7.4 | 72 | 0.6 | 83.4 | 11.6 | 18.8 | 19.2 | 0.0004 |
| RBD Σ[S/CO] | 21 | 1.8 | 44.7 | 6.9 | 10.4 | 10.6 | 72 | 0.9 | 44.3 | 9.7 | 12.7 | 10.8 | 0.0067 |
| S Σ[S/CO] | 21 | 1.2 | 39.9 | 9.8 | 13.4 | 10.7 | 72 | 0.8 | 187.0 | 19.5 | 37.2 | 42.5 | 0.2217 |
| N/RBD/S Σ[S/CO] | 21 | 4.8 | 116.9 | 27.4 | 30.0 | 25.9 | 72 | 3.4 | 290.6 | 45.2 | 68.6 | 68.6 | 0.0091 |

**Note*: N, nucleocapsid; RBD, receptor binding domain; S, spike; ECD, Spike ectodomain; Σ[S/CO], sum of signal to cut-off ratio; Gen, GenScript; NAC, Native Antigen Company; Sino, Sino Biological.

†Wilcoxon-Mann-Whitney test.

**Supplementary Table 7. Median fluorescence intensity (MFI) cut-off values derived using 324 pre-pandemic saliva samples, collected prior to January, 2019.***

| Antigen | N† | cut-off‡ | $\log_{10}$(cut-off) |
|---|---|---|---|
| NAC N | 265 | 810 MFI | 2.91 MFI |
| Gen N | 265 | 1174 MFI | 3.07 MFI |
| Sino RBD | 265 | 334 MFI | 2.52 MFI |
| Gen RBD | 265 | 447 MFI | 2.65 MFI |
| Mt. Sinai RBD | 265 | 156 MFI | 2.19 MFI |
| Sino ECD | 265 | 1123 MFI | 3.05 MFI |
| Mt. Sinai S | 265 | 922 MFI | 2.96 MFI |
| N Σ[S/CO] | 265 | 1.73 | 0.24 |
| RBD Σ[S/CO] | 265 | 2.65 | 0.42 |
| S Σ[S/CO] | 265 | 1.89 | 0.28 |
| N/RBD/S Σ[S/CO] | 265 | 6.00 | 0.78 |

**Note*: N, nucleocapsid; RBD, receptor binding domain; S, spike; ECD, Spike ectodomain; Σ[S/CO], sum of signal to cut-off ratio; Gen, GenScript; NAC, Native Antigen Company; Sino, Sino Biological

†pre-pandemic negative saliva samples collected before January 2019

‡Mean + 3 standard deviations; units are median fluorescence intensity (MFI) for individual antigens and Σ[S/CO] values are unitless
